# The Adipo-B Index as a Novel Integrator of Glycemic and Lipid Homeostasis: a Multiple-Therapy Validation Study

**DOI:** 10.64898/2026.02.16.26346332

**Authors:** Eiji Kutoh, Alexandra N. Kuto

**Author notes:** Address for correspondence: Eiji Kutoh, M.D., Ph.D., Biomedical Center, 1-5-8-613 Komatsugawa, Edogawa-ku 132-0034, Tokyo, Japan.

## Abstract

**Objective:** To introduce and evaluate the clinical utility of the “adipo-B index” as a novel metric of the adipose tissue-pancreatic beta cell axis. To our knowledge, no prior clinical metric has integrated adipose tissue insulin resistance and pancreatic beta-cell function into a single index applicable across therapeutic classes.

**Methods:** Treatment-naïve subjects with T2DM received monotherapy with modified traditional diet for diabetes (MJDD, n=61), canagliflozin (n=67), pioglitazone (n=54), or sitagliptin (n=63). Correlations between the baseline and changes in adipo-IR or adipo-B and clinical parameters were analyzed. This is a prospective, non-randomized observational study.

**Results:** At baseline, among all the subjects, adipo-B significantly correlated with FBG, HbA1c and non-HDL-C, while adipo-IR did not. At 3 months, across all therapeutic strategies, significant negative correlations were observed between the changes in (Δ)adipo-B and baseline adipo-B. By contrast, in MJDD, canagliflozin and pioglitazone, significant negative correlations were seen between Δadipo-IR and baseline adipo-IR, while with sitagliptin, no correlations were noted. Δadipo-B, but not Δadipo-IR, correlated with the improvements of glycemic (FBG, HbA1c) and lipid (non-HDL-C) parameters across all these therapies. While significant correlations were seen between Δadipo-B and Δadipo-IR with MJDD, pioglitazone and sitagliptin, canagliflozin uniquely “decoupled” this axis. With sitagliptin and pioglitazone, adipo-B improved despite weight gain.

**Conclusion:** The adipo-B index is a superior indicator of systemic metabolic status and therapeutic response and could serve as a useful tool for precision therapy for diabetes.

## Introduction

Conventional biomarkers and indices for T2DM, such as glycemic parameters (FBG, HbA1c), insulin resistance (HOMA-R, adipo-IR), beta-cell function (HOMA-B), or diabetic dyslipidemia (non-HDL-C) are often analyzed in isolation (1, 2). However, metabolic health is not determined by a single tissue, but by the coordinated modulation of multiple tissues. To date, no validated metric has successfully quantified this crosstalk across different therapeutic classes.

Adipo-IR is frequently utilized to quantify the failure of insulin to suppress the lipolytic release of free fatty acids (FFAs) in adipose tissue, a primary driver of systemic lipotoxicity, which, in turn, deteriorates pancreatic beta-cell function (3, 4). The systemic impact of adipo-IR is ultimately determined by the capacity of the pancreatic beta-cell to compensate for this lipotoxic load. Thus, examining these metrics individually provides an incomplete picture of metabolic status. A high degree of adipo-IR may be clinically silent if beta-cell function remains robust enough to neutralize the resulting lipotoxic load. Overt hyperglycemia and dyslipidemia typically emerge only when this metabolic mismatch exceeds the ability of the pancreas to compensate. Despite this, a unified metric that integrates these two traditional parameters has remained elusive in clinical practice.

In this study, to investigate whether the control of some diabetic parameters is driven by adipose tissue status in isolation or by the balance between adipose-derived stress and pancreatic response, we introduce the adipo-B index (adipo-IR divided by HOMA-B) as a novel integrator of systemic metabolic status. This composite metric serves to normalize the metabolic load against the metabolic work being performed by the pancreas.

By analyzing a cohort of treatment-naïve subjects with T2DM across four major distinct therapeutic arms in monotherapy-tight diet (MJDD), sodium glucose co-transporter (SGLT)-2 inhibitor, thiazolidinedione (TZD) and dipeptidyl peptidase (DPP)-4 inhibitor-we aimed to: 1) validate the index against standard glycemic and lipid markers, 2) compare its predictive power to that of adipo-IR alone, and 3) investigate the unique decoupling effects of different pharmacological classes on the adipose tissue-pancreas axis.

The findings in this study suggest that the adipo-B index offers a superior, high-resolution view of therapeutic response that remains invisible to structural metrics such as BMI. We propose the adipo-B index as a functional measure of this “adipose tissue-pancreas” cross-talk and this novel index may represent a paradigm shift in metabolic phenotyping.

## Subjects and Methods

### Subjects

The study was conducted in accordance with the Declaration of Helsinki and was approved by the institutional review board (IRB) of Gyoda General Hospital and Kumagaya Surgery Hospital. Written informed consent was obtained and recorded electronically. This project is an extension of the registered study that is listed in the UMIN database (ID: UMIN000006860). Newly diagnosed, treatment-naive subjects with T2DM without any emergent conditions (e.g. ketoacidosis) were enrolled according to Japan Diabetes Society criteria, as described previously (5). Exclusion criteria: renal impairment (creatinine ≥1.5 mg/dL), hepatic dysfunction (AST/ALT ≥70 IU/L), heart disease, severe hypertension (>160/100 mmHg), T1DM, or pregnancy. The subjects were recruited from Gyoda General Hospital, Kumagaya Surgery Hospital (Saitama, Japan), and other affiliated sites of the first author (EK) between March 2019 and December 2025. The dropout subjects were excluded from analysis. The final cohort included 245 subjects, assigned to either a modified Japanese diet for diabetes (MJDD, n=61), canagliflozin (n=67), pioglitazone (n=54) or sitagliptin (n=63). No other interventions were used. The subjects were not strictly randomized; this is a comparison of four observational cohorts. The details of MJDD have been published (5). Canagliflozin was given at 50 mg/day for women and 100 mg/day for men. Pioglitazone was given 15 mg/day for women and 30 mg/day for men. Sitagliptin was given at 25 mg for women and 50 mg for men. The canagliflozin, pioglitazone and sitagliptin groups also followed standard dietary advice.

### Laboratory Measurements

The primary endpoint was change in adipo-IR and adipo-B after 3 months of treatment. The secondary endpoints included FBG, HbA1c, insulin, FFA, HOMA-R, HOMA-B and BMI. Fasting blood was collected before breakfast. HbA1c and FBG were measured monthly; insulin at baseline and 3 months (Abbott Japan). In some suspected subjects, anti-GAD antibodies were checked to exclude T1DM (Mitsubishi BML).

Calculations (1, 2, 5)

HOMA-R = (insulin × FBG) / 405

adipo-IR = FFA × Insulin

HOMA-B = (insulin × 360) / (FBG – 63)

adipo-B=adipo-IR / HOMA-B

### Data Analysis

Changes were calculated as baseline to 3-month differences. Paired Student’s t-tests assessed intra-group changes; simple regression evaluated correlations. The results are presented as mean ± SD. Significance was set at p<0.05; p-values between 0.05 and 0.1 were considered to represent a trend toward significance. The analyses were performed using PAST software (https://www.nhm.uio.no/english/research/resources/past/index.html) from University of Oslo

## Results

### Baseline Correlations of Adipo-IR and Adipo-B with Clinical Parameters

Simple regression analyses were performed between the baseline levels of adipo-IR or adipo-B and various diabetes-related parameters in all enrolled subjects. As shown in Table 1A, significant positive correlations were observed between the baseline adipo-IR and insulin, FFA, HOMA-R, HOMA-B and BMI; however, no associations were found with FBG, HbA1c, or non-HDL-C. Conversely, as shown in Table 1B, baseline levels of the adipo-B index were significantly positively correlated with FBG, HbA1c, FFA and non-HDL-C, and were negatively correlated with HOMA-B. No correlations were observed between adipo-B and adipo-IR, insulin, HOMA-R or BMI.

**Table 1.** Correlations between baseline values of adipo-IR/adipo-B and diabetic parameters (all subjects) Simple regression analyses were performed between the baseline values of adipo-IR or adipo-B and some diabetic parameters for all subjects (n=245). Panel A) adipo-IR and diabetic parameters Panel B) adipo-B and diabetic parameters

| baseline adipo-IR vs. baseline | R | p-values |
| --- | --- | --- |
| adipo-B | 0.175 | n.s. |
| FBG | -0.074 | n.s. |
| HbA1c | -0.091 | n.s. |
| insulin | 0.841 | <0.00001 |
| FFA | 0.36 | <0.00001 |
| HOMA-R | 0.807 | <0.00001 |
| HOMA-B | 0.668 | <0.00001 |
| non-HDL-C | 0.127 | n.s. |
| BMI | 0.625 | <0.00001 |

B
| baseline adipo-B vs. baseline | R | p-values |
| --- | --- | --- |
| adipo-R | 0.175 | n.s. |
| FBG | 0.754 | <0.00001 |
| HbA1c | 0.617 | <0.00001 |
| insulin | -0.124 | n.s. |
| FFA | 0.65 | <0.00001 |
| HOMA-R | 0.155 | n.s. |
| HOMA-B | -0.378 | <0.00001 |
| non-HDL-C | 0.32 | <0.00001 |
| BMI | -0.057 | n.s. |

### Changes in Diabetic Parameters Across Four Therapeutic Strategies

The subjects received monotherapy as follows:

**MJDD (Table 2A):** Significant reductions in adipo-IR, adipo-B, FBG, HbA1c, insulin, FFA, HOMA-R, non-HDL-C, and BMI were observed, alongside significant increases in HOMA-B.

**Table 2.** Changes in diabetic parameters across different therapeutic strategies. Paired Student’s t-tests were used to compare the changes in the indicated parameters at baseline and 3 months treatment. The results are expressed as the mean ± SD (standard deviation). A) MJDD B) canagliflozin C) pioglitazone D) sitagliptin

| n=61 | baseline | 3 months | p-values |
| --- | --- | --- | --- |
| age | 51.6±14 |  |  |
| adipo-IR | 5.66±4.09 | 4.64±3.63 | <0.02 |
| adipo-B | 0.246±0.154 | 0.203±0.145 | <0.02 |
| FBG (mg/dl) | 182.7±45.5 | 170.9±54.4 | <0.04 |
| HbA1c (%) | 8.97±1.28 | 8.04±1.55 | <0.00001 |
| insulin (μU/ml) | 7.57±4.94 | 6.72±3.94 | <0.02 |
| FFA (mEq/L) | 0.780±0.360 | 0.659±0.227 | <0.01 |
| HOMA-R | 3.41±2.39 | 2.75±1.69 | <0.02 |
| HOMA-B | 25.75±18.88 | 29.35±24.01 | <0.05 |
| non-HDL-C (mg/dl) | 161.2±42.4 | 153.5±32.2 | <0.05 |
| BMI | 25.83±4.88 | 25.06±4.58 | <0.00001 |

| n=67 | baseline | 3 months | p-values |
| --- | --- | --- | --- |
| age | 53.9±13.0 |  |  |
| adipo-IR | 5.72±4.31 | 5.27±4.13 | 0.091 |
| adipo-B | 0.299±0.169 | 0.195±0.108 | <000001 |
| FBG (mg/dl) | 197.0±61.9 | 146.2±37.0 | <0.00001 |
| HbA1c (%) | 10.07±2.52 | 8.23±1.85 | <0.00001 |
| insulin (μU/ml) | 7.13±5.64 | 6.36±5.05 | <0.02 |
| FFA (mEq/L) | 0.818±0.292 | 0.875±0.363 | n.s. |
| HOMA-R | 3.37±2.84 | 2.30±1.87 | <0.00001 |
| HOMA-B | 24.69±24.63 | 31.48±27.2 | <0.0002 |
| non-HDL-C (mg/dl) | 168.0±45.5 | 166.8±41.7 | n.s. |
| BMI | 26.13±5.24 | 25.65±5.19 | <0.00001 |

| n=54 | baseline | 3 months | p-values |
| --- | --- | --- | --- |
| age | 54.4±11.1 |  |  |
| adipo-IR | 4.86±4.94 | 3.43±4.31 | <0.006 |
| adipo-B | 0.337±0.200 | 0.211±0.151 | <0.00001 |
| FBG (mg/dl) | 223.4±61.8 | 192.6±73.3 | <0.0006 |
| HbA1c (%) | 10.0±11.87 | 9.09±2.04 | <0.00001 |
| insulin (μU/ml) | 6.71±6.90 | 6.17±6.12 | n.s. |
| FFA (mEq/L) | 0.743±0.354 | 0.582±0.254 | <0.004 |
| HOMA-R | 3.42±2.87 | 2.73±2.48 | <0.01 |
| HOMA-B | 19.96±27.94 | 24.66±32.03 | 0.075 |
| non-HDL-C (mg/dl) | 158.0±43.9 | 149.1±38.7 | <0.05 |
| BMI | 24.32±5.49 | 24.69±5.78 | <0.003 |

| n=63 | baseline | 3 months | p-values |
| --- | --- | --- | --- |
| age | 55.8±11.7 |  |  |
| adipo-IR | 5.63±4.18 | 5.87±6.37 | n.s. |
| adipo-B | 0.381±0.256 | 0.290±0.257 | <0.0002 |
| FBG (mg/dl) | 217.4±62.9 | 191.3±76.6 | <0.0002 |
| HbA1c (%) | 10.10±2.26 | 8.23±2.18 | <0.00001 |
| insulin (μU/ml) | 6.58±4.23 | 7.53±6.04 | <0.05 |
| FFA (mEq/L) | 0.859±0.339 | 0.752±0.326 | <0.02 |
| HOMA-R | 3.50±2.73 | 3.70±4.01 | n.s. |
| HOMA-B | 18.75±13.01 | 26.47±19.47 | <0.0002 |
| non-HDL-C (mg/dl) | 169.6±43.1 | 165.9±39.6 | n.s. |
| BMI | 24.54±4.53 | 24.82±4.43 | <0.03 |

**Canagliflozin (Table 2B):** Significant reductions in adipo-B, FBG, HbA1c, insulin, HOMA-R, and BMI were observed, with significant increases in HOMA-B. Adipo-IR showed a trend toward a reduction, while FFA and non-HDL-C remained unchanged.

**Pioglitazone (Table 2C):** Significant reductions in adipo-IR, adipo-B, FBG, HbA1c, FFA, HOMA-R, and non-HDL-C were observed, while BMI significantly increased. HOMA-B showed a tendency to increase, and insulin levels remained stable.

**Sitagliptin (Table 2D):** Significant reductions in adipo-B, FBG, HbA1c, insulin, and FFA were observed, while HOMA-B and BMI significantly increased. No changes were noted for adipo-IR, HOMA-R, or non-HDL-C.

### Correlation Between Changes (Δ) and Baseline Levels of Adipo-IR and Adipo-B

Simple regression analyses evaluated the relationship between the changes (Δ) in adipo-IR or Δadipo-B and their respective baseline levels.

1. With MJDD, canagliflozin and pioglitazone, significant negative correlations were observed between Δadipo-IR and baseline adipo-IR while with sitagliptin, no correlations were noted (Fig. 1A, 1B, 1C, 1D).
2. With all therapeutic strategies, significant negative correlations were seen between Δadipo-B and the baseline levels of adipo-B (Fig. 2A, 2B, 2C, 2D).

**Figure 1.**
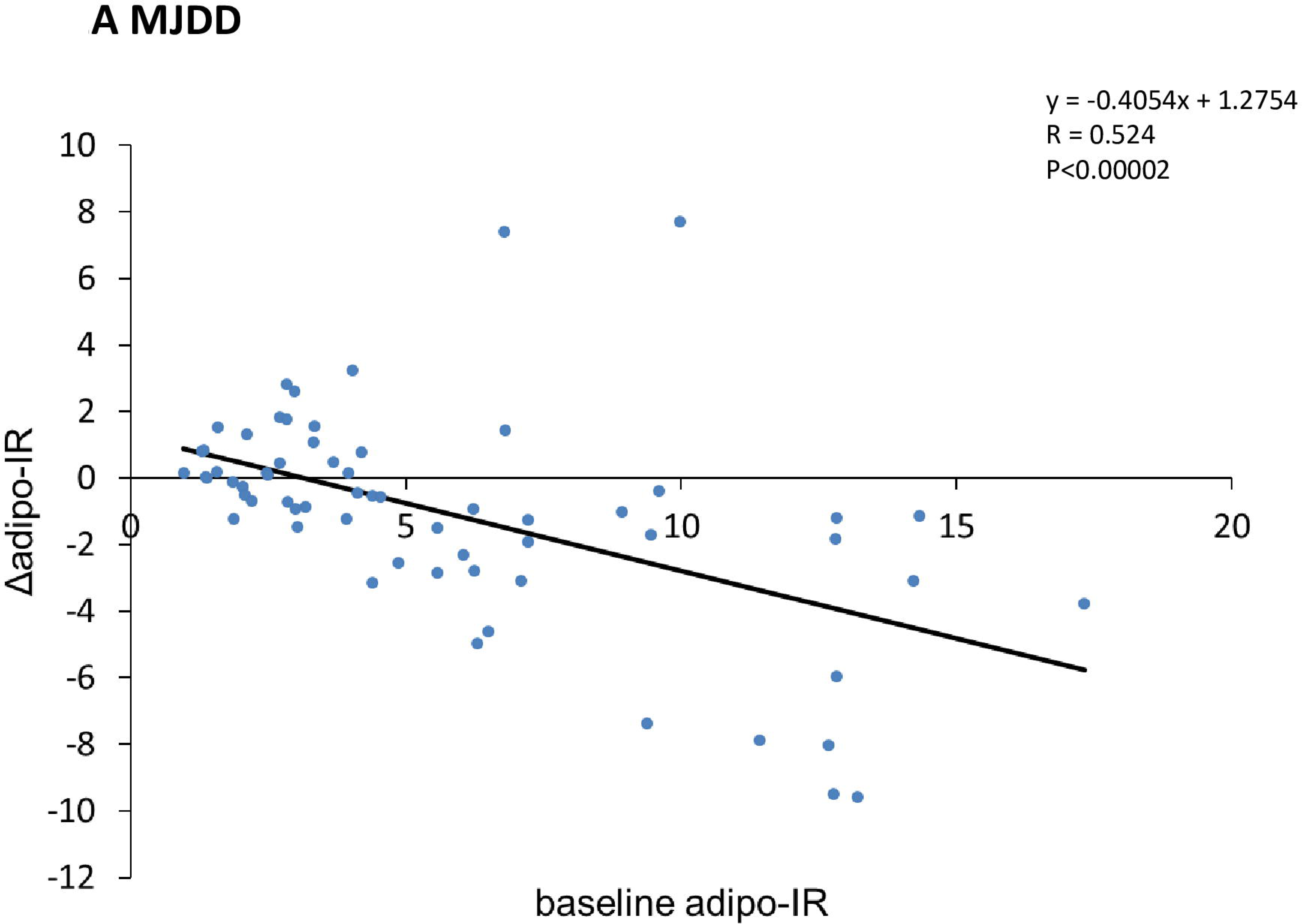
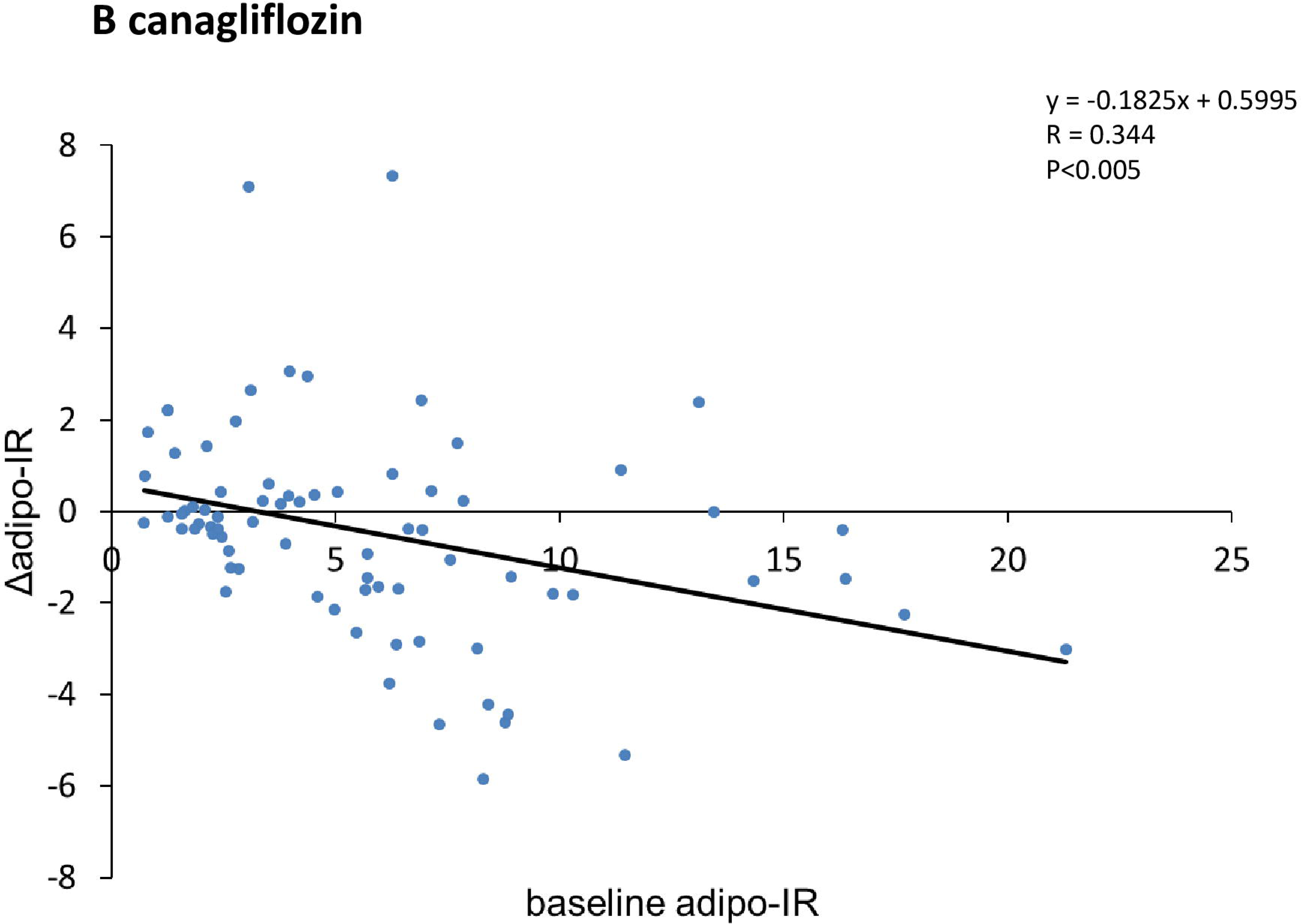
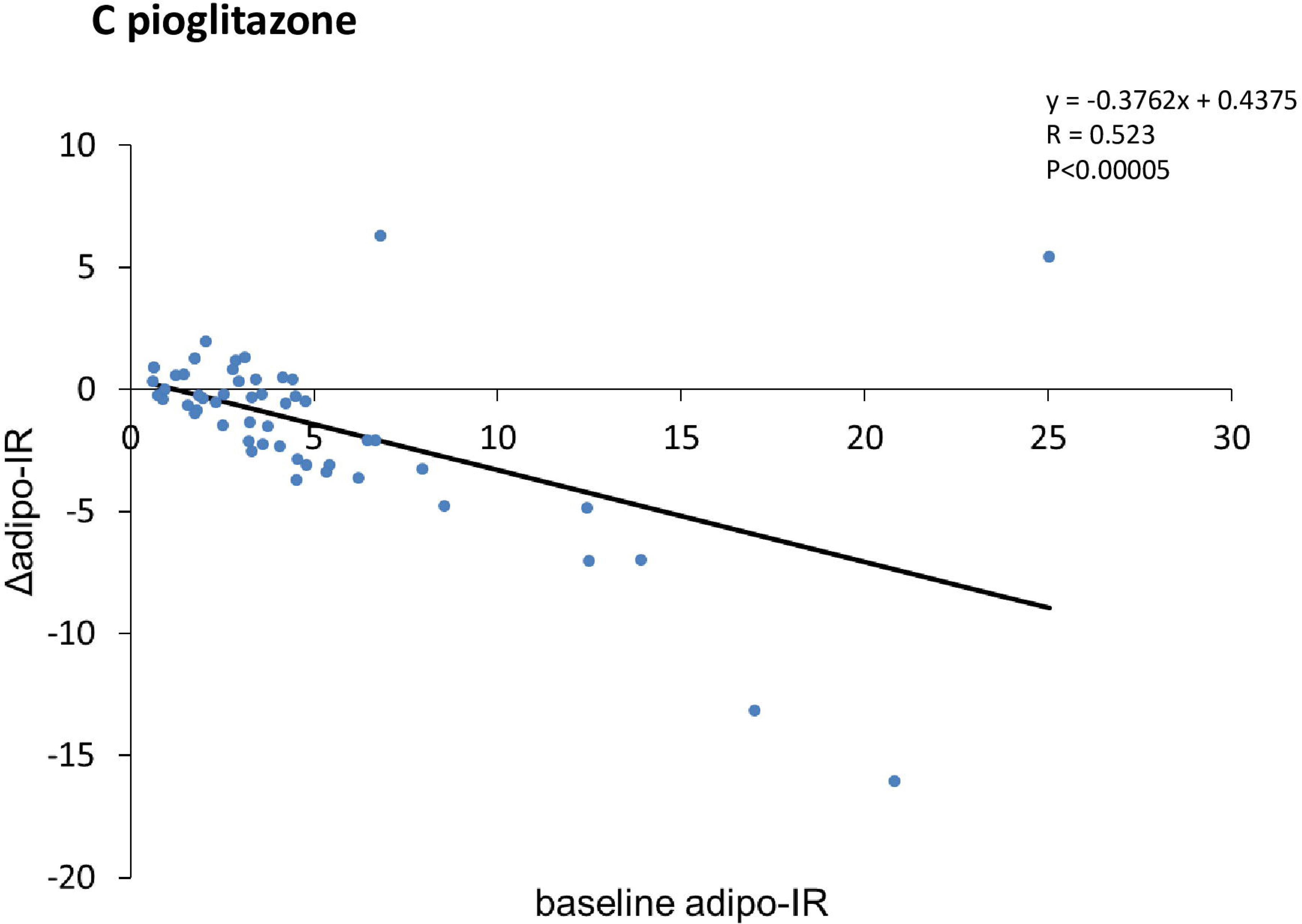
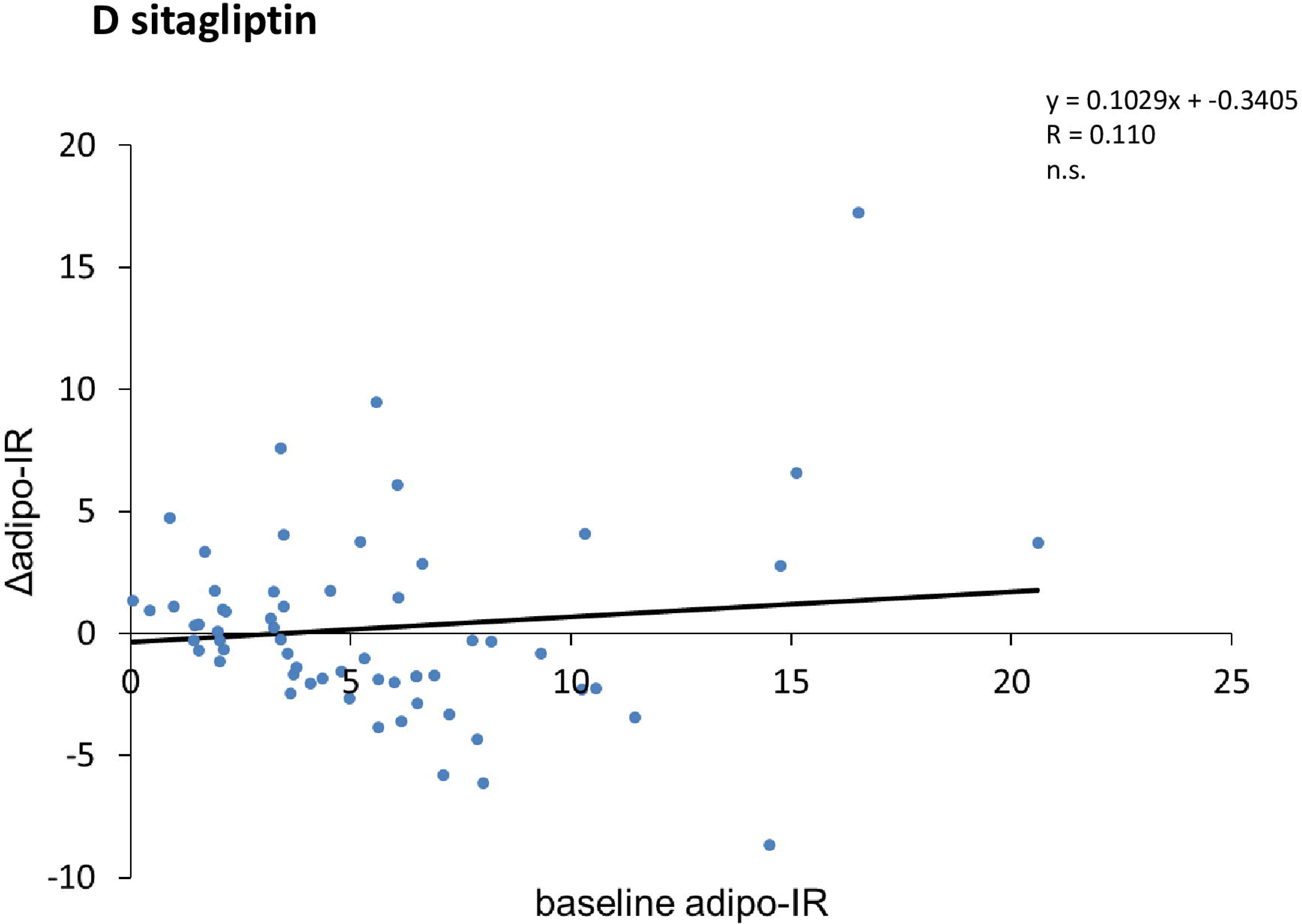
Baseline-dependent regulation of adipo-IR. Simple regression analyses were performed between the changes of (Δ)adipo-IR and the baseline adipo-IR Panel A) MJDD Panel B) canagliflozin Panel C) pioglitazone Panel D) sitagliptin

**Figure 2.**
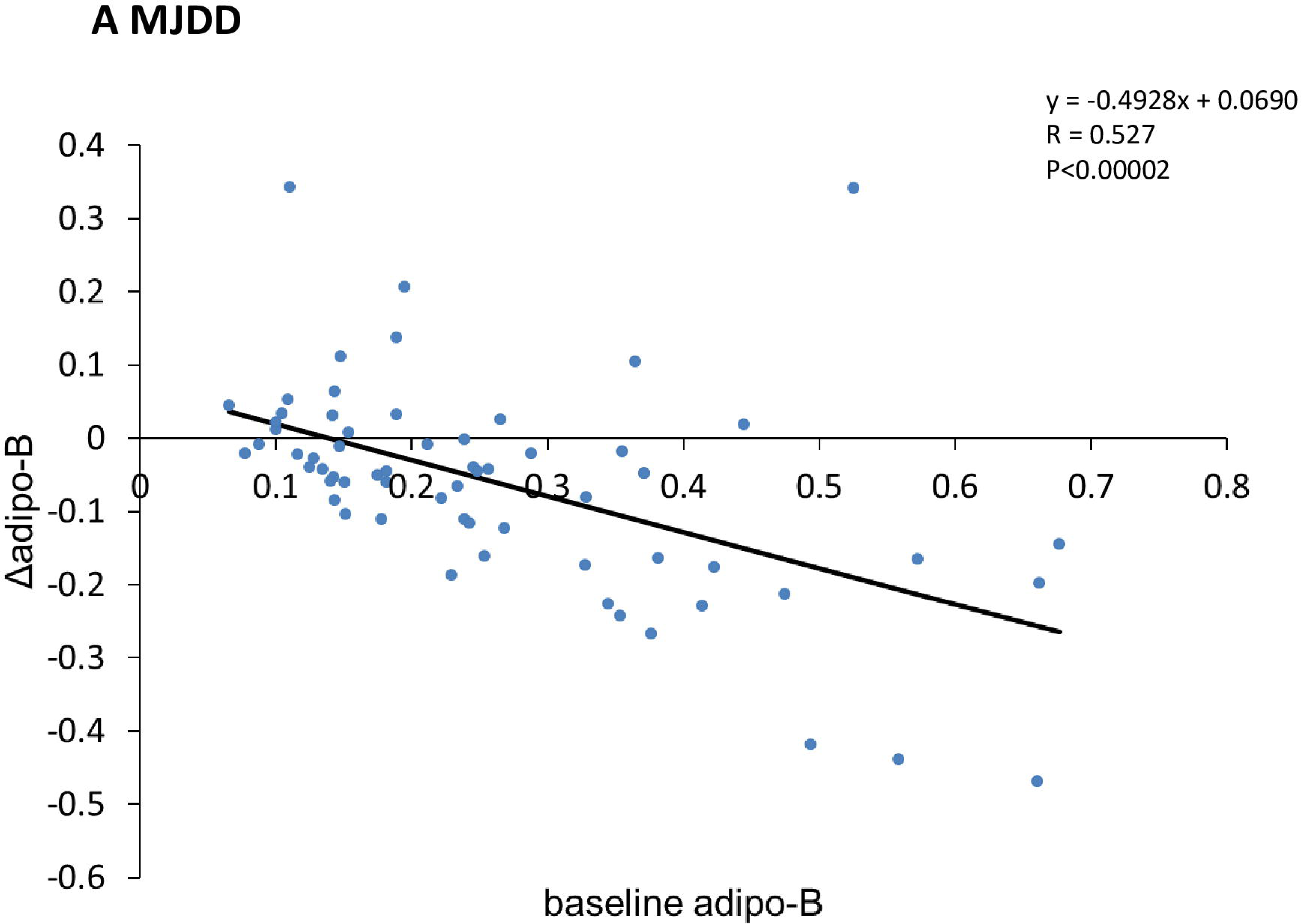
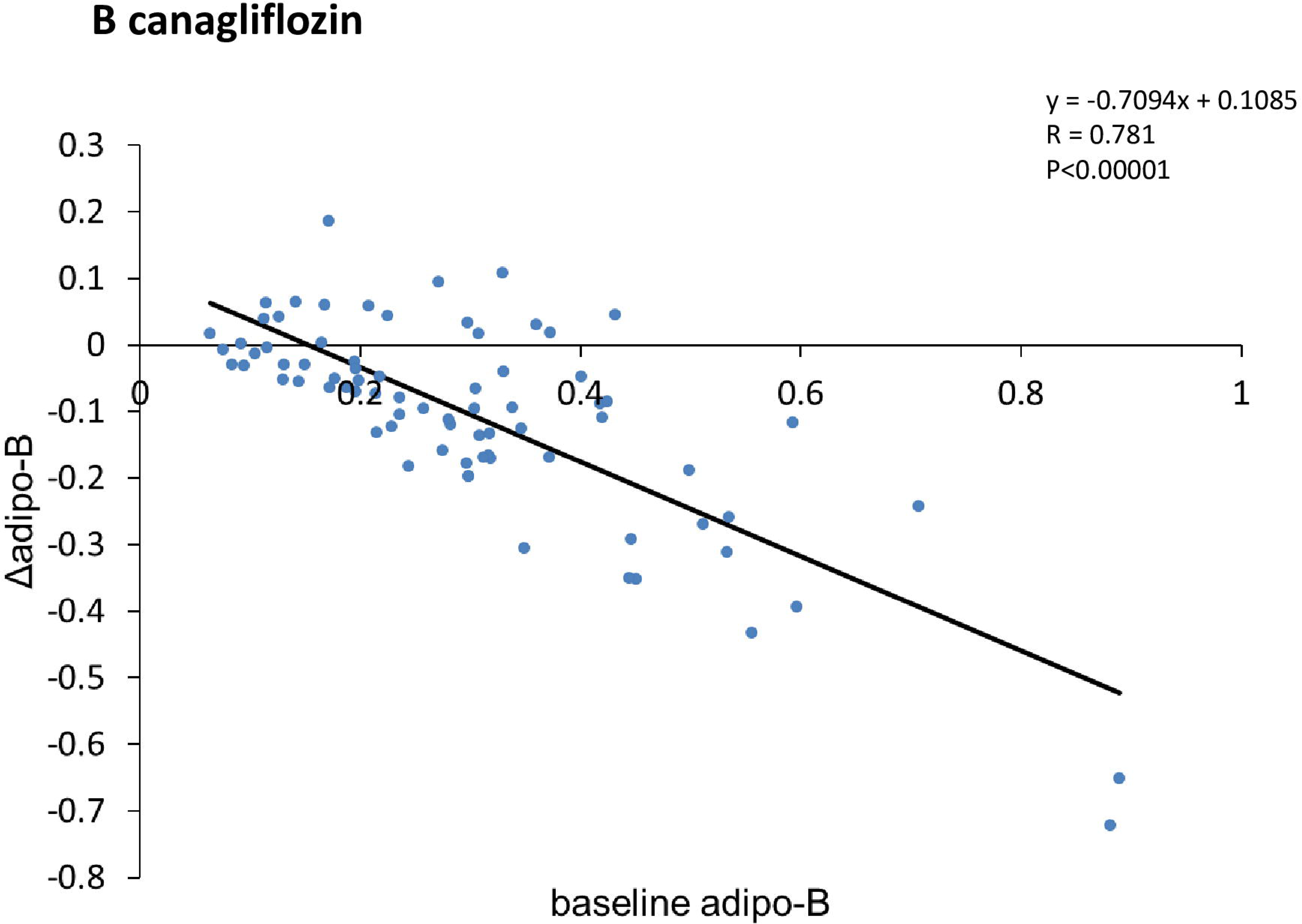
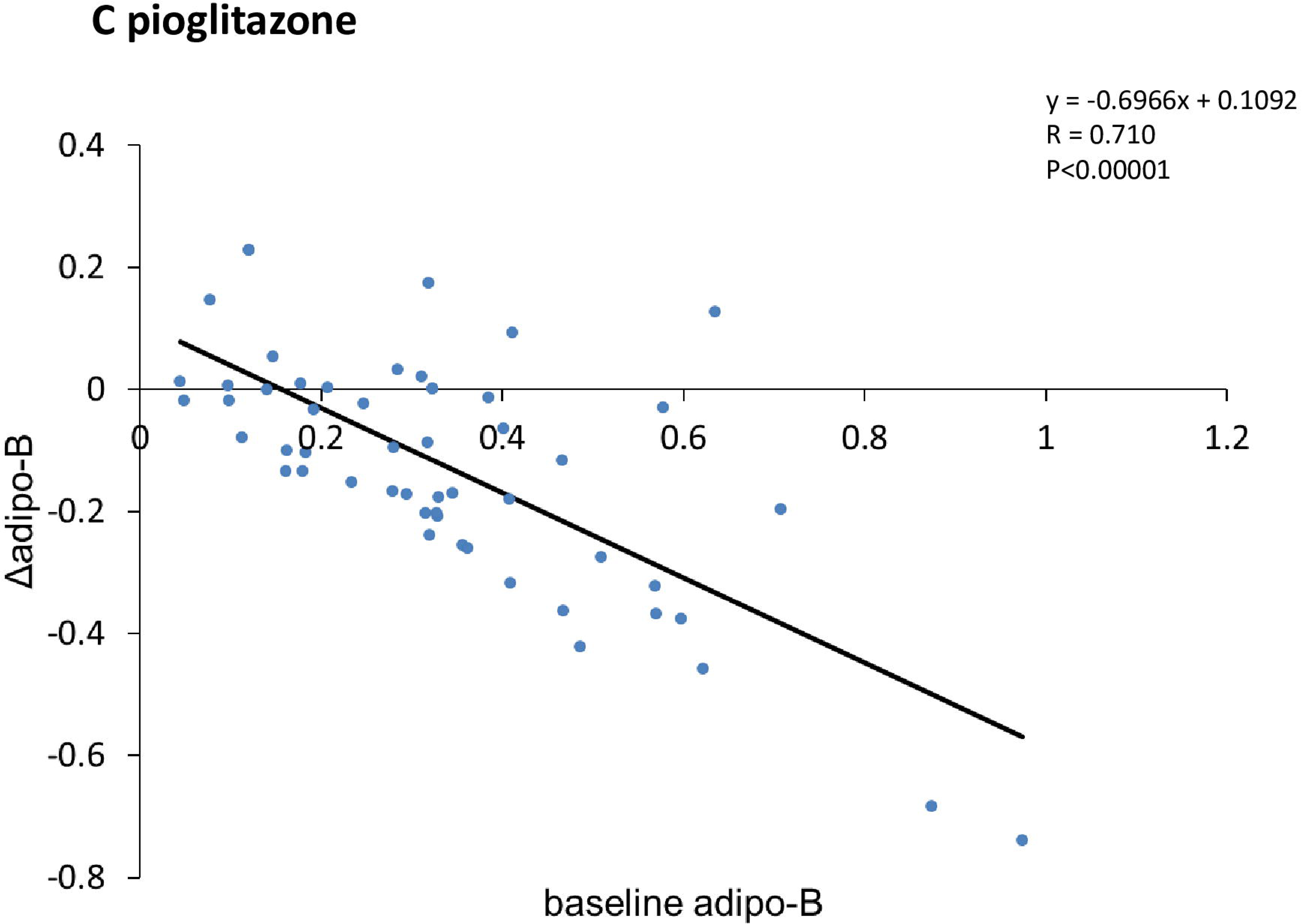
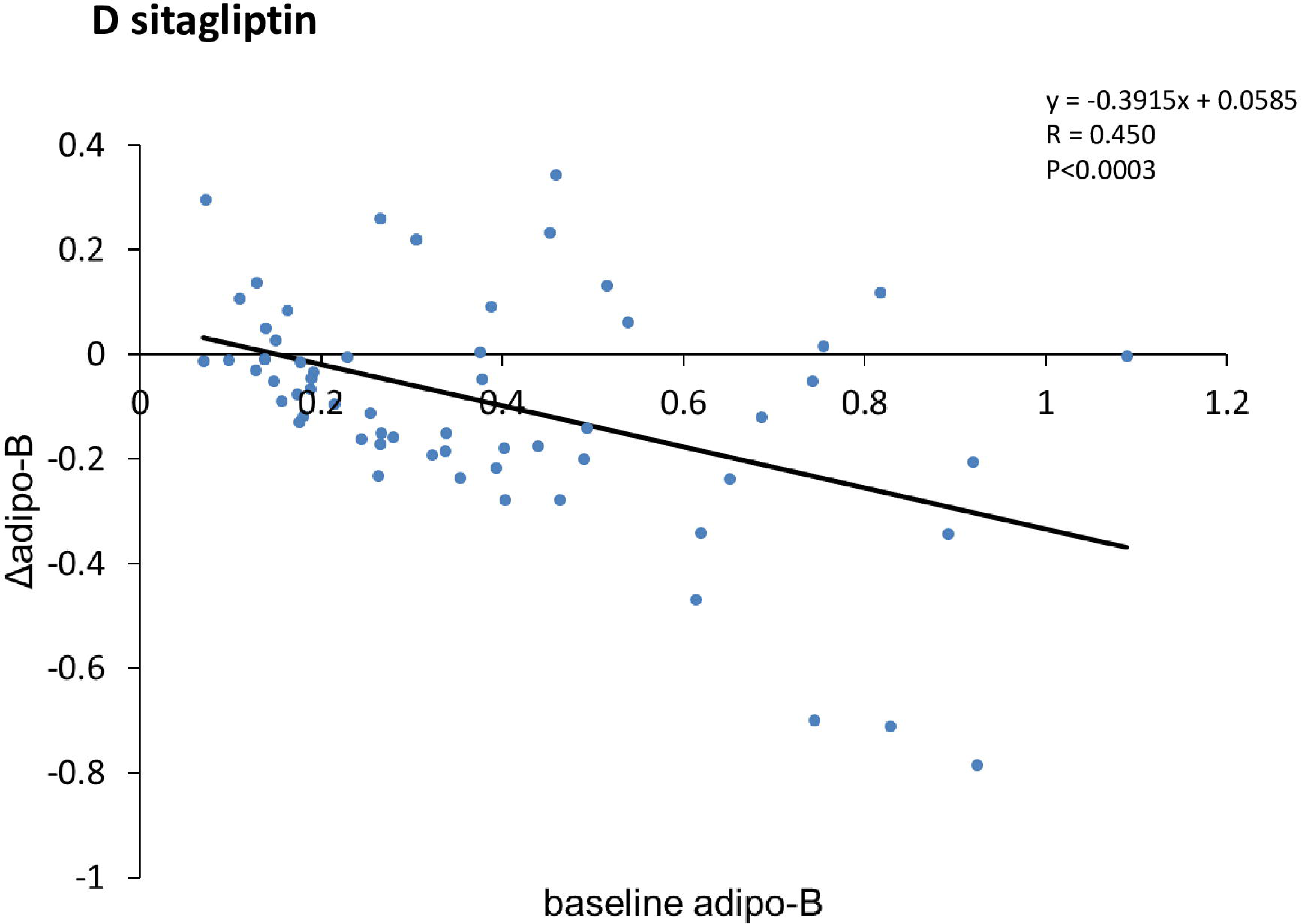
Baseline dependent regulation of adipo-B. Simple regression analyses were performed between the changes of (Δ)adipo-B and the baseline adipo-B Panel A) MJDD Panel B) canagliflozin Panel C) pioglitazone Panel D) sitagliptin

### Correlations Between Changes (**Δ**) in Adipo-IR and Diabetic Parameters

Simple regression analyses evaluated the relationship between the changes (Δ) in adipo-IR and those of diabetes-related parameters:

**MJDD, pioglitazone, and sitagliptin (Tables 3A, 3C, 3D):** Δadipo-IR significantly correlated with Δadipo-B, Δinsulin, and ΔFFA across these groups. Notably, Δadipo-IR did not correlate with ΔFBG, ΔHbA1c, or ΔBMI.

**Canagliflozin (Table 3B):** Δadipo-IR correlated with ΔFFA, Δinsulin, and ΔHOMA-R/B, but uniquely showed no correlation with Δadipo-B.

**Table 3.**
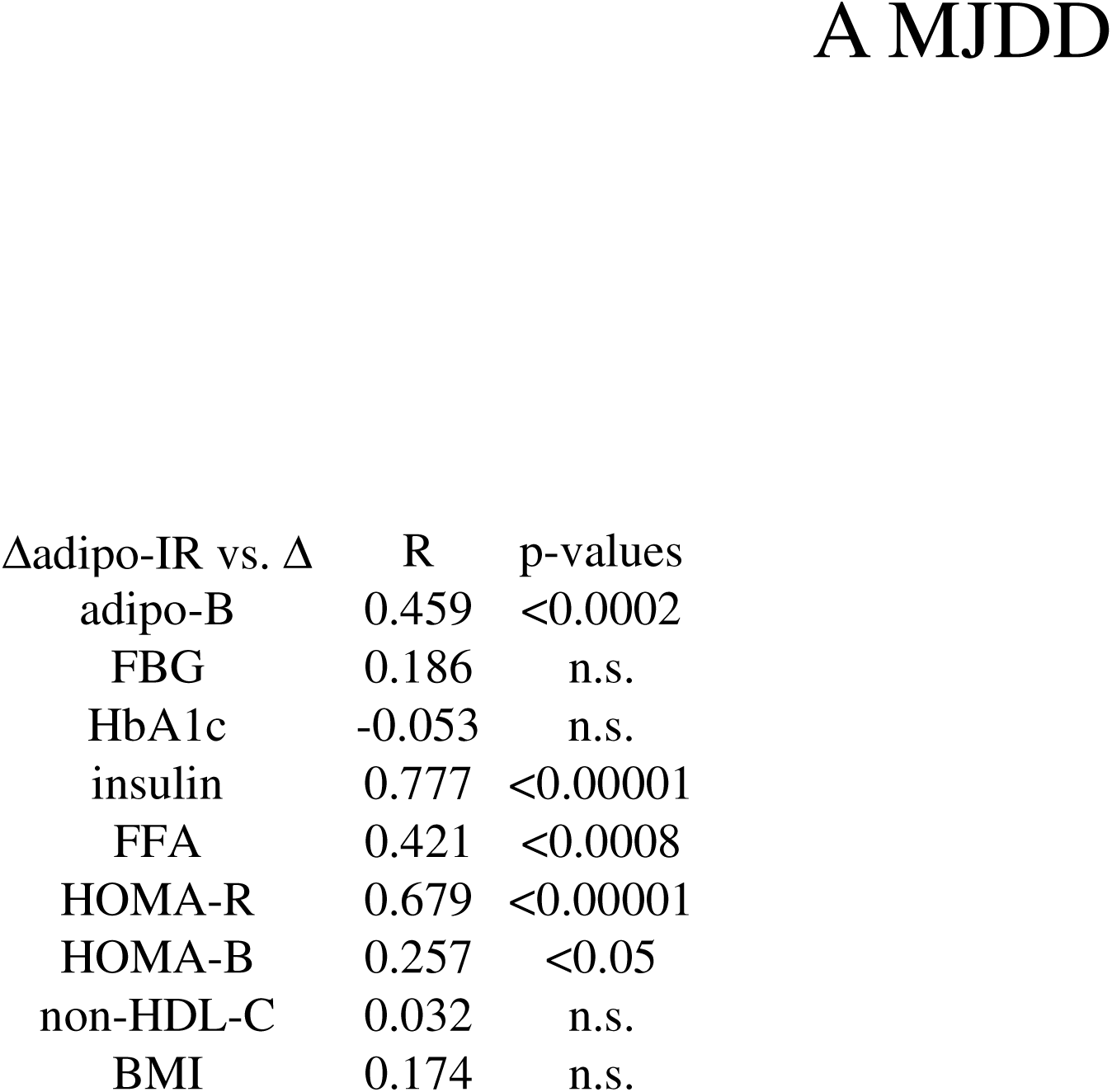
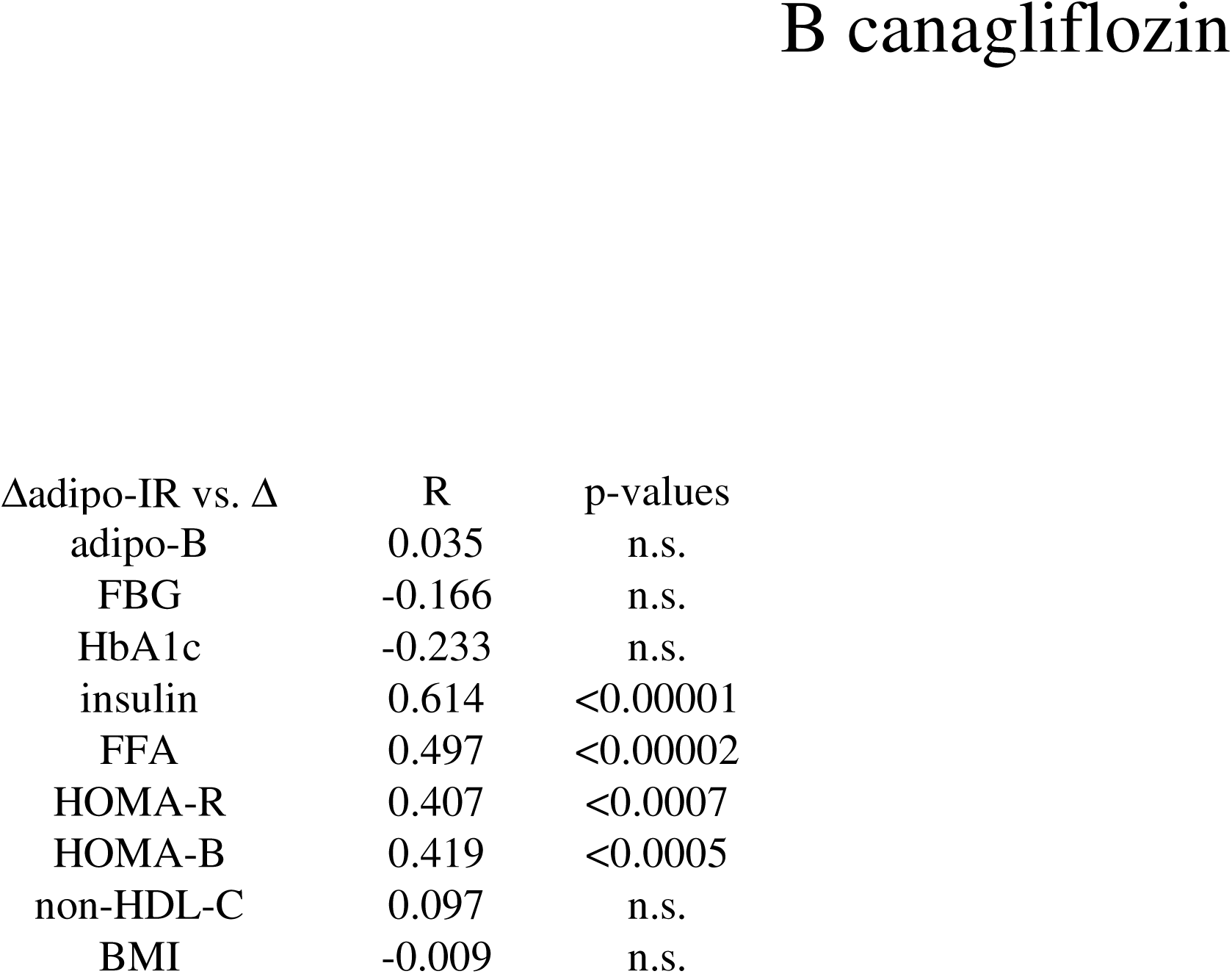
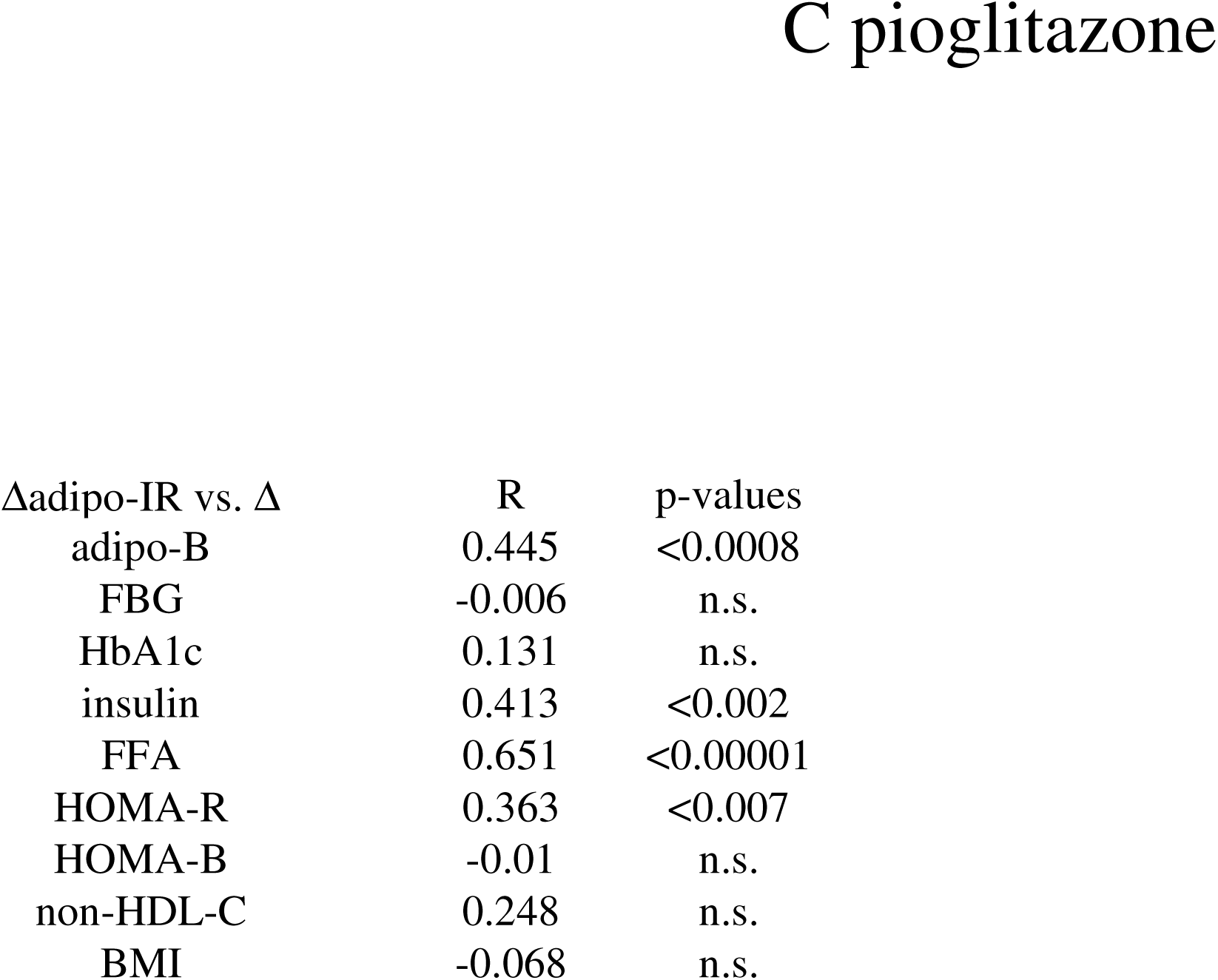
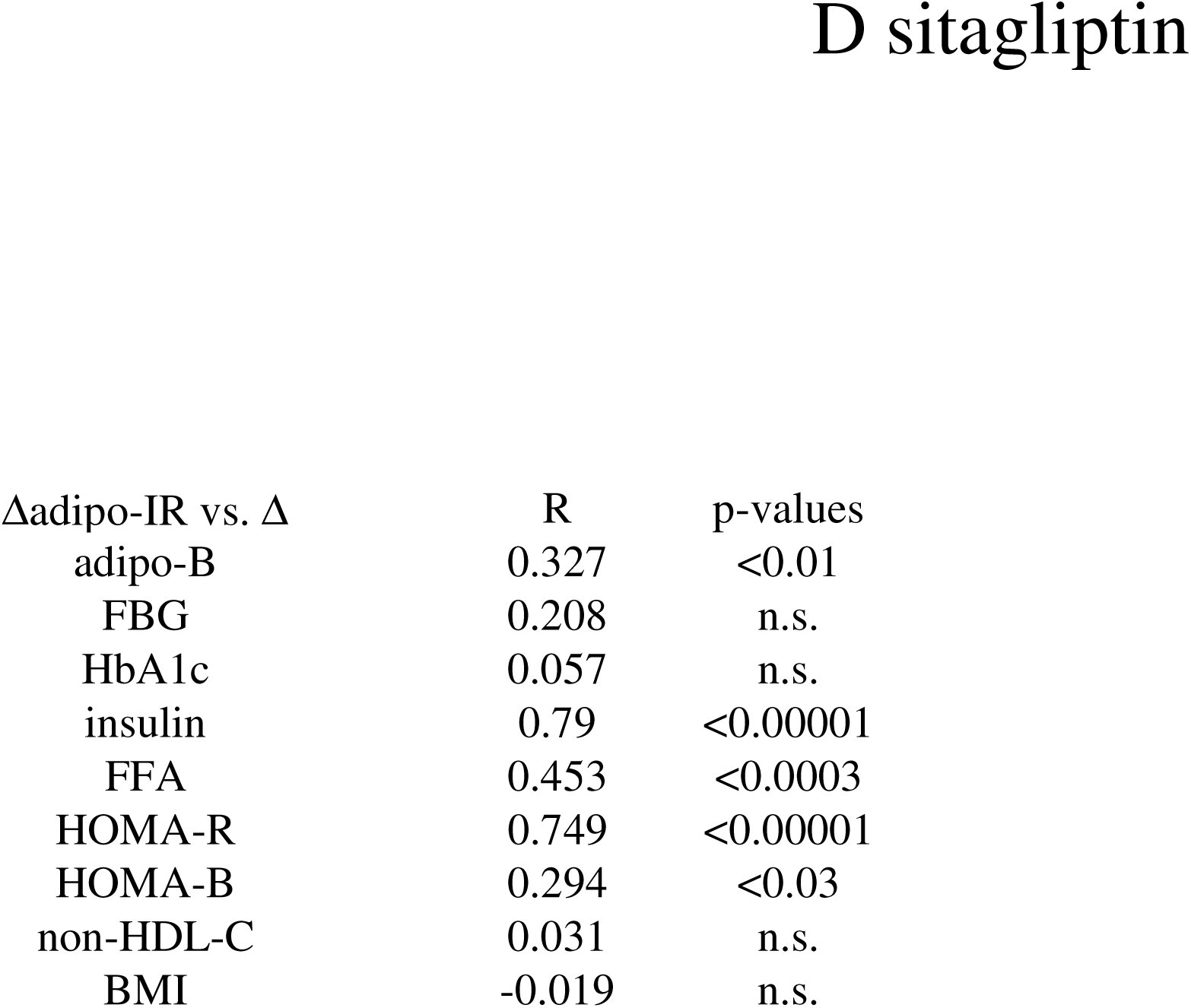
Correlations between the changes (Δ) in adipo-IR and diabetic parameters. Simple regression analyses evaluated the relationship between the changes (Δ) in adipo-IR and those of diabetes-related parameters: A) MJDD B) canagliflozin C) pioglitazone D) sitagliptin

| Δadipo-IR vs. $\Delta$ | R | p-values |
| --- | --- | --- |
| adipo-B | 0.459 | <0.0002 |
| FBG | 0.186 | n.s. |
| HbA1c | -0.053 | n.s. |
| insulin | 0.777 | <0.00001 |
| FFA | 0.421 | <0.0008 |
| HOMA-R | 0.679 | <0.00001 |
| HOMA-B | 0.257 | <0.05 |
| non-HDL-C | 0.032 | n.s. |
| BMI | 0.174 | n.s. |
| adipo-B | 0.035 | n.s. |
| FBG | -0.166 | n.s. |
| HbA1c | -0.233 | n.s. |
| insulin | 0.614 | <0.00001 |
| FFA | 0.497 | <0.00002 |
| HOMA-R | 0.407 | <0.0007 |
| HOMA-B | 0.419 | <0.0005 |
| non-HDL-C | 0.097 | n.s. |
| BMI | -0.009 | n.s. |
| adipo-B | 0.445 | <0.0008 |
| FBG | -0.006 | n.s. |
| HbA1c | 0.131 | n.s. |
| insulin | 0.413 | <0.002 |
| FFA | 0.651 | <0.00001 |
| HOMA-R | 0.363 | <0.007 |
| HOMA-B | -0.01 | n.s. |
| non-HDL-C | 0.248 | n.s. |
| BMI | -0.068 | n.s. |
| adipo-B | 0.327 | <0.01 |
| FBG | 0.208 | n.s. |
| HbA1c | 0.057 | n.s. |
| insulin | 0.79 | <0.00001 |
| FFA | 0.453 | <0.0003 |
| HOMA-R | 0.749 | <0.00001 |
| HOMA-B | 0.294 | <0.03 |
| non-HDL-C | 0.031 | n.s. |
| BMI | -0.019 | n.s. |

### Correlations Between Changes (**Δ**) in Adipo-B and Diabetic Parameters

**Cross-Group Efficacy (Tables 4A, 4B, 4C, 4D):** In all four groups, Δadipo-B significantly, positively correlated with ΔFBG, ΔHbA1c, and Δnon-HDL-C.

**Weight Divergence:** In the MJDD and canagliflozin groups, Δadipo-B did not correlate with ΔBMI (Tables 4A, 4B). However, in the pioglitazone and sitagliptin groups, significant negative correlations were observed between Δadipo-B and ΔBMI (Tables 4C, 4D).

## Discussion

The present study introduces and validates the adipo-B index as a novel, integrated biomarker of the adipose tissue-pancreatic beta-cell axis. Our findings demonstrate that while adipo-IR serves as a localized measure of adipose tissue dysfunction, it is the balance between this lipotoxic stress and pancreatic compensatory capacity-quantified by the adipo-B index-that dictates systemic glycemic and lipid homeostasis.

### The Superiority of Integrated Metrics over Isolated Biomarkers

A fundamental observation in the baseline data was the lack of correlation between adipo-IR and primary glycemic or lipid markers (HbA1c, FBG, non-HDL-C, Table 1A). This dissociation suggests that adipose tissue insulin resistance, while a driver of metabolic stress, does not lead to overt hyperglycemia as long as the pancreatic beta-cells can maintain hyperinsulinemic compensation. In contrast, the adipo-B index showed robust correlations with HbA1c, FBG, and non-HDL-C (Table 1B). By incorporating HOMA-B into the denominator, the index successfully identifies the tipping point where pancreatic reserve fails to neutralize the flux of FFAs, leading to systemic metabolic collapse.

### Baseline Predictors of Change: The Primacy of the Adipo-B Index

The analysis of baseline characteristics as predictors of therapeutic response reveals a critical distinction between adipose-specific (adipo-IR) and axis-integrated (adipo-B) metrics. As shown in Fig. 1A, 1B and 1C, for MJDD, canagliflozin, and pioglitazone, the magnitude of improvement in adipo-IR was directly proportional to the baseline severity of adipose tissue insulin resistance (dysfunction). However, sitagliptin efficacy was notably independent of baseline adipo-IR (Fig. 1D), suggesting a mechanistic ceiling in its ability to modulate adipocyte-specific insulin resistance. Crucially, however, the adipo-B index functioned as a universal predictor of response across all four therapeutic arms (Fig. 2A, 2B, 2C, 2D). Regardless of the mechanism(s) of action-whether strict diet, renal, adipose-targeted, or incretin-based-those with the highest baseline adipo-B index experienced the most significant improvements. This suggests that while individual drugs may have blind spots regarding adipose tissue, the adipo-B index captures a fundamental state of metabolic instability that is amenable to intervention across all major classes of therapy of T2DM.

### Mechanistic Divergence: The Decoupling Effect of SGLT-2 Inhibition

Our therapeutic analysis revealed a striking mechanistic distinction between canagliflozin and other interventions. In the MJDD, pioglitazone, and sitagliptin arms, changes in adipo-B were tightly tethered to changes in adipo-IR (Table 3A, 3C, 3D). This implies that these therapies improve the adipose tissue-pancreas axis primarily by reducing the load on the pancreas or enhancing the sensitivity of adipocytes to insulin. However, canagliflozin fundamentally decoupled this relationship (Table 3B). Despite only a marginal trend in adipo-IR reduction, canagliflozin achieved significant improvements in both adipo-B and HbA1c (Table 2B). We propose that canagliflozin (and probably other SGLT-2 inhibitors as well) acts as a renal safety valve, providing a glucose-clearance pathway that is entirely independent of the adipose tissue-pancreas feedback loop. This “metabolic resilience” makes SGLT-2 inhibitors an essential tool for those with refractory adipose tissue resistance who have exhausted their pancreatic capacity.

### Metabolic Quality vs. Structural Quantity

The divergence between BMI and adipo-B in the sitagliptin and pioglitazone groups provides critical evidence for the clinical utility of this index. While both drugs were associated with weight gain in this cohort (Table 2C, 2D)-a finding consistent with certain Japanese populations (6, 7)-the adipo-B index consistently improved. This suggests that the index captures metabolic quality (the functional state of lipid-glucose handling) rather than adipose quantity. The negative correlation between Δadipo-B and ΔBMI with pioglitazone and sitagliptin (Table 4C, 4D) indicates that these drugs may promote the expansion of “healthy” subcutaneous fat that acts as a metabolic sink, thereby reducing the lipotoxic pressure on the beta-cells despite an increase in total body mass.

**Table 4.**
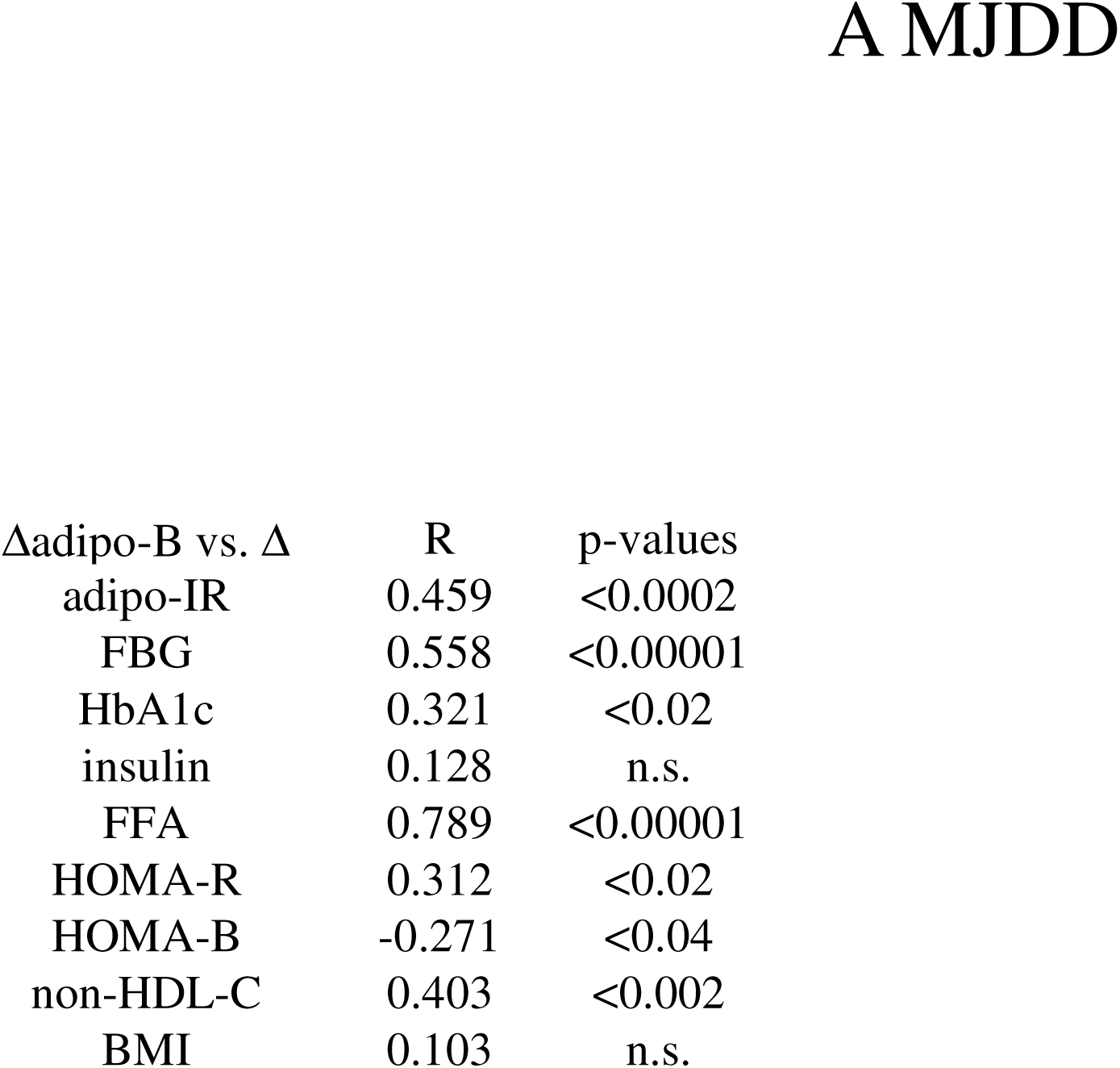
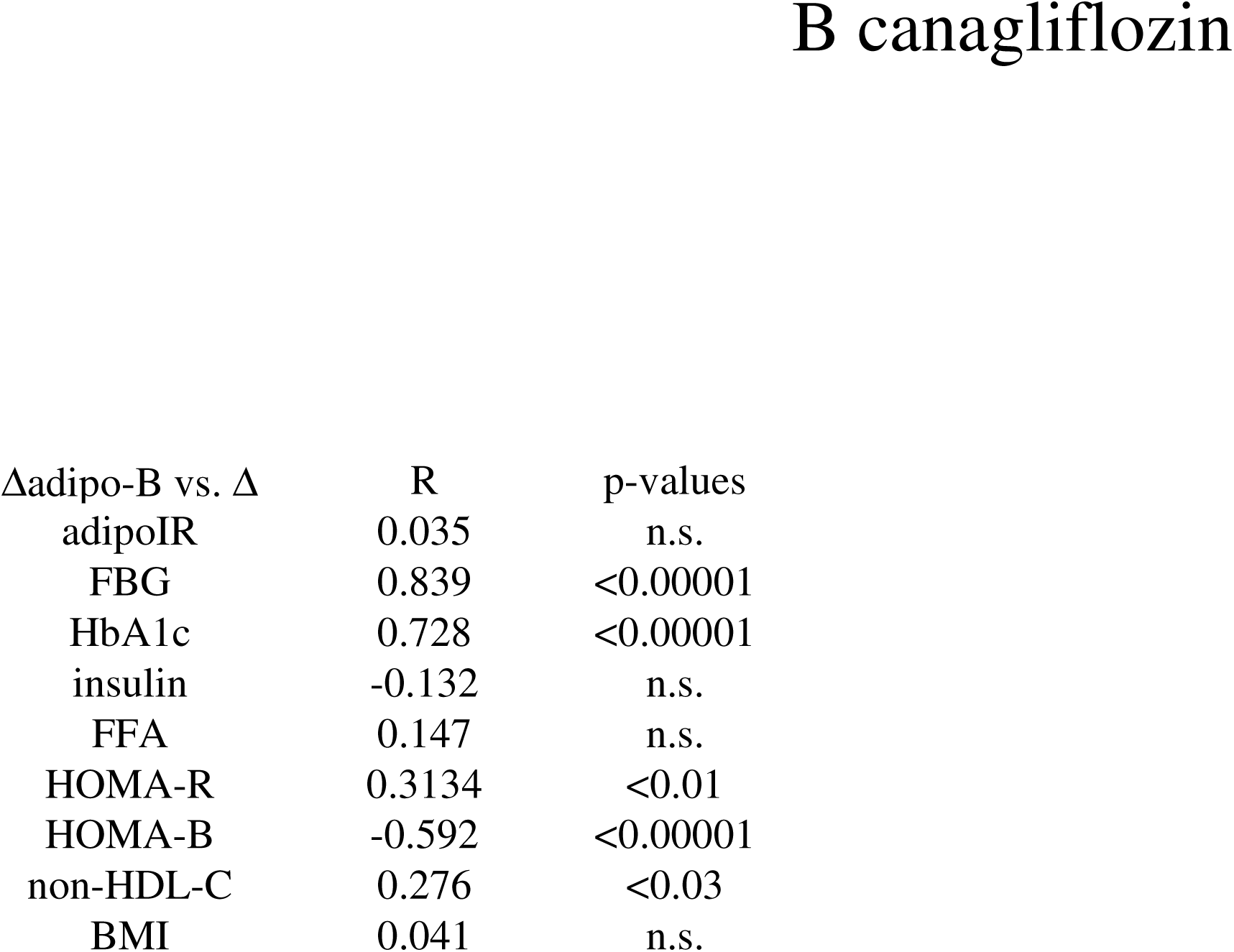
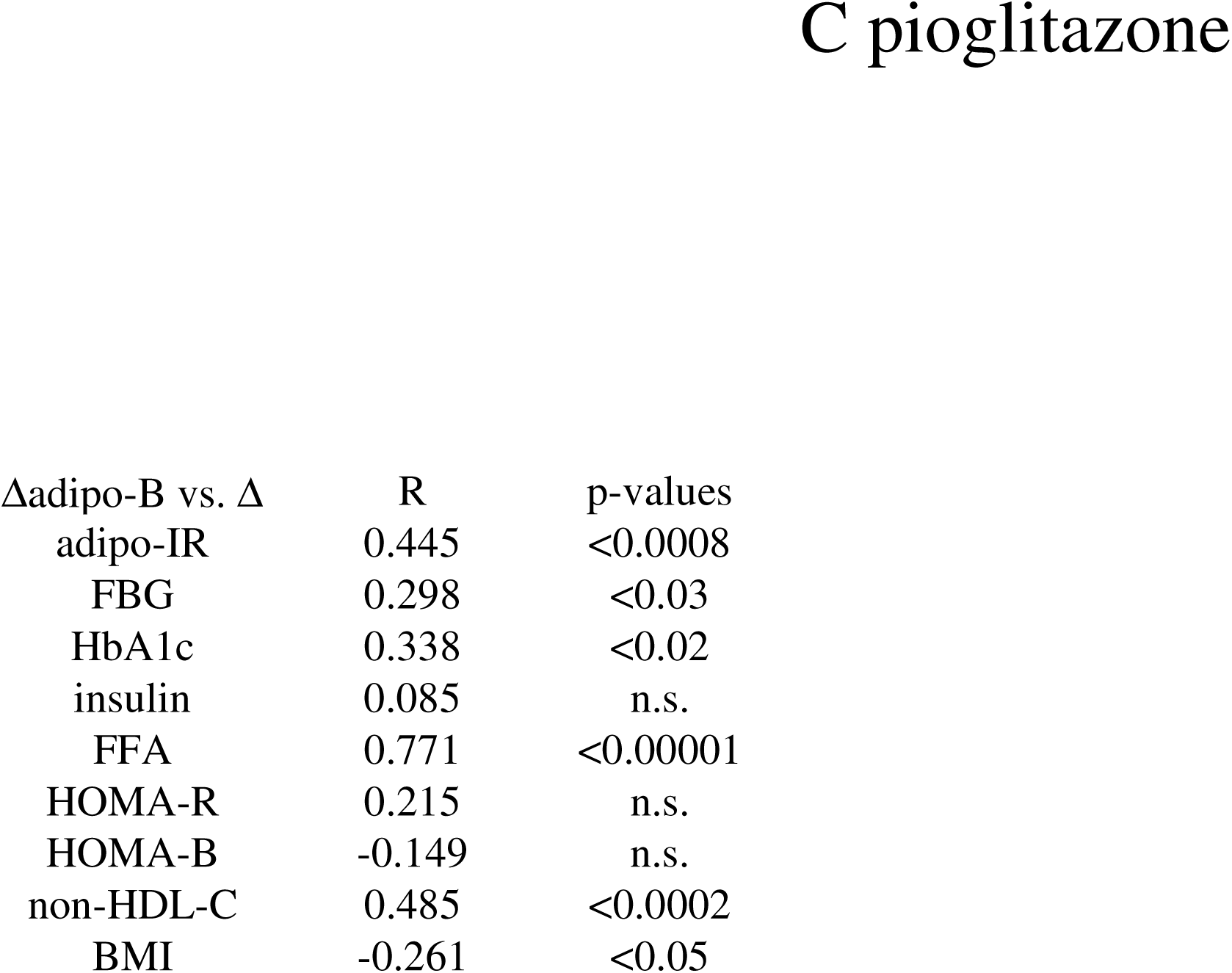
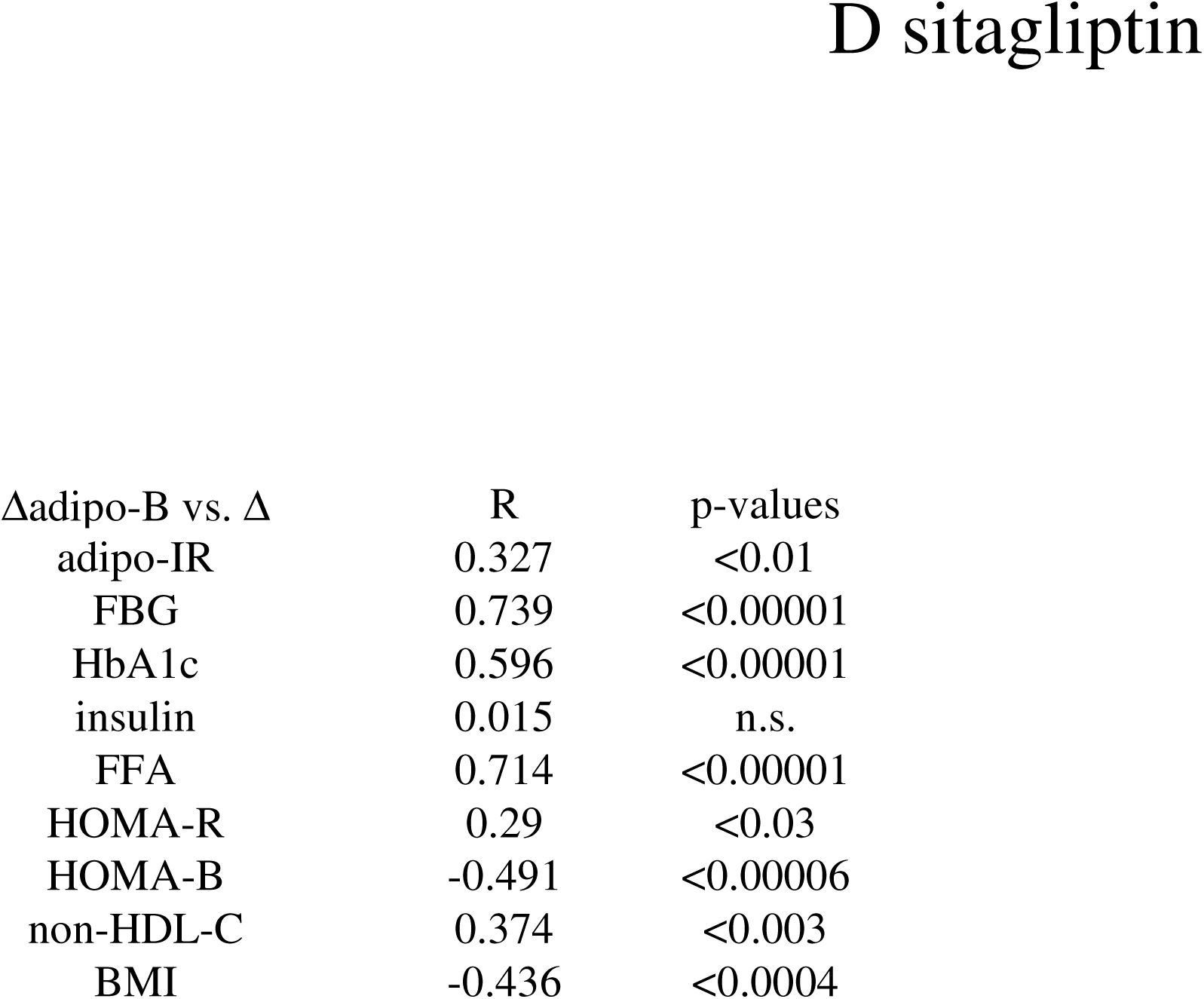
Correlations between the changes (Δ) in adipo-B and diabetic parameters. Simple regression analyses evaluated the relationship between the changes (Δ) in adipo-B and those of diabetes-related parameters: A) MJDD B) canagliflozin C) pioglitazone D) sitagliptin

| Δadipo-B vs. $\Delta$ | R | p-values |
| --- | --- | --- |
| adipo-IR | 0.459 | <0.0002 |
| FBG | 0.558 | <0.00001 |
| HbA1c | 0.321 | <0.02 |
| insulin | 0.128 | n.s. |
| FFA | 0.789 | <0.00001 |
| HOMA-R | 0.312 | <0.02 |
| HOMA-B | -0.271 | <0.04 |
| non-HDL-C | 0.403 | <0.002 |
| BMI | 0.103 | n.s. |
| adipoIR | 0.035 | n.s. |
| FBG | 0.839 | <0.00001 |
| HbA1c | 0.728 | <0.00001 |
| insulin | -0.132 | n.s. |
| FFA | 0.147 | n.s. |
| HOMA-R | 0.3134 | <0.01 |
| HOMA-B | -0.592 | <0.00001 |
| non-HDL-C | 0.276 | <0.03 |
| BMI | 0.041 | n.s. |
| adipo-IR | 0.445 | <0.0008 |
| FBG | 0.298 | <0.03 |
| HbA1c | 0.338 | <0.02 |
| insulin | 0.085 | n.s. |
| FFA | 0.771 | <0.00001 |
| HOMA-R | 0.215 | n.s. |
| HOMA-B | -0.149 | n.s. |
| non-HDL-C | 0.485 | <0.0002 |
| BMI | -0.261 | <0.05 |
| adipo-IR | 0.327 | <0.01 |
| FBG | 0.739 | <0.00001 |
| HbA1c | 0.596 | <0.00001 |
| insulin | 0.015 | n.s. |
| FFA | 0.714 | <0.00001 |
| HOMA-R | 0.29 | <0.03 |
| HOMA-B | -0.491 | <0.00006 |
| non-HDL-C | 0.374 | <0.003 |
| BMI | -0.436 | <0.0004 |

### Clinical Implications for Precision Therapy for Diabetes

The adipo-B index might offer a practical framework for therapeutic triage. Those with high adipo-B driven by failing HOMA-B are likely to be non-responders to incretin-based therapies like DPP-4 inhibitors, which rely on endogenous pancreatic reserve. For these individuals, initiating axis-independent therapies like SGLT-2 inhibitors early in the treatment algorithm may be necessary to bypass the failing adipose tissue-pancreas axis and prevent further lipotoxic damage.

### Strengths and Limitations

#### Strengths

A major strength of this study is the prospective, multi-arm design, which allowed for a head-to-head comparison of the adipo-B index across four distinct therapeutic modalities. By including both lifestyle interventions and pharmacological agents with diverse mechanisms of action, we were able to demonstrate the versatility of the index in capturing varied metabolic responses. Furthermore, the inclusion of treatment-naïve subjects eliminated the confounding influence of other medications, providing a clear window into the primary physiological effects of each intervention on the adipose tissue-pancreas axis. The strong correlation of the index with non-HDL cholesterol and BMI (Table 4A, 4B, 4C, 4D) further reinforces its utility as a comprehensive marker of systemic metabolic health rather than a narrow indicator of hyperglycemia.

### Limitations

Despite these strengths, several limitations warrant consideration. First, as a preliminary observational, non-randomized study, the sample size within each individual treatment arm was small, which may limit the generalizability of the findings to broader or more ethnically diverse populations. Second, the follow-up period was focused on short-to-medium-term metabolic shifts; long-term durability of the adipo-B index as a predictor of cardiovascular outcomes or diabetes remission remains to be established. Finally, while HOMA-B and adipo-IR are validated surrogate markers, they do not provide the same level of precision as hyperinsulinemic-euglycemic clamps or intravenous glucose tolerance tests. Larger, multi-center validation cohorts are required to confirm these findings and to establish standardized clinical cutoff values for the adipo-B index across different stages of T2DM progression.

## Conclusions

The adipo-B Index provides a practical, calculation-based tool for precision diabetology. It allows physicians to identify “metabolic responders and non-responders” early and select therapies (like SGLT-2 inhibitors) that operate outside the failing adipose tissue-pancreas axis. This research might provide both a new physiological framework and a useful diagnostic tool for the global diabetes research community.

## Data Availability

All data produced in the present study available upon reasonable request to the authors.

## Conflicts of Interest

The authors declare that no conflicts of interest exist regarding this manuscript.

## CRediT authorship contribution statement

E.K. and A.N.K. participated in the design of the study and acquisition of the data, performed the statistical analysis, and drafted the manuscript. Writing, review and editing: E.K. and A.N.K. All authors have read and agreed to the published version of the manuscript.

## Funding

This research received no external funding.

## Informed Consent Statement

Informed consent was obtained from all patients and stored in the electronic medical record system.

## Data Availability Statement

The data that support the findings of this study are available from the corresponding author (E.K.) upon reasonable request.

## Acknowledgments

The authors express gratitude to Asuka Wada, Midori Akiyama, Rumi Kurihara, Nozomi Urishibara, Rumiko Okada for their insightful discussions and valuable feedbacks. The authors also appreciate Keiko Saido-Ozawa for her administrative support.

## Abbreviations

T2DM: type 2 diabetes
BMI: body mass index
adipo-IR: adipose tissue insulin resistance
FBG: fasting blood glucose
Non-HDL-C: non-high density lipoprotein cholesterol
HOMA-R: homeostasis model assessment-R
HOMA-B: homeostasis model assessment-B

## Notes

### Competing Interest Statement

The authors have declared no competing interest.

### Funding Statement

This study did not receive any funding.

### Author Declarations

The study was conducted in accordance with the Declaration of Helsinki and was approved by the institutional review board (IRB) of Gyoda General Hospital and Kumagaya Surgery Hospital.

### Summary of Updates

BMI in Table A and B were mislabeled. The manuscript of these parts are corrected accordingly (abstract and result section).

